# A high-throughput amplicon screen for somatic *UBA1* variants in Cytopenic and Giant Cell Arteritis cohorts

**DOI:** 10.1101/2021.12.08.21266919

**Authors:** James A. Poulter, Alesia Khan, Stephen Martin, Mark Grey, Bosko Andjelic, Emma Nga, Diana I.D. Triantafyllopoulou, Paul Evans, Louise Sorensen, Sarah L. Mackie, UKGCA Consortium, Ann W. Morgan, Catherine Cargo, Sinisa Savic

**Author notes:** See Appendix 1.

## Abstract

Somatic mutations in the gene encoding the major E1 ubiquitin ligase, UBA1, were recently identified as a cause of VEXAS, a late-onset acquired auto-inflammatory syndrome. Differential diagnoses for patients subsequently found to have VEXAS include relapsing polychondritis, Sweet’s syndrome, myelodysplastic syndrome (MDS), giant cell arteritis (GCA) and undifferentiated systemic autoinflammatory disease (uSAID). We therefore sought to screen DNA from individuals with a non-diagnostic cytopenia or GCA, for known VEXAS-associated mutations. To this end, we developed a multiplexed UBA1 amplicon sequencing assay, allowing quick screening of large cohorts while also providing sufficient sequencing depth to identify somatic mutations to an allele frequency <1%. Using this assay, we screened genomic DNA from 612 males diagnosed with GCA, and bone marrow DNA from 1,055 cases with an undiagnosed cytopenia. No GCA cases were found to have *UBA1* mutations, however 4 different mutations in the cytopenic cohort were identified in 7 individuals. Furthermore, we describe a female case identified in the screen with a *UBA1* mutation and all VEXAS-associated phenotypes, but without Monosomy X. Our study suggests that, despite the overlap in clinical features, VEXAS is rarely misdiagnosed as GCA, but identified in 1.0% of males with an undiagnosed cytopenia. The identification of a *UBA1* variant in a female case adds further evidence that VEXAS should not be ruled out as a differential diagnosis in females with VEXAS-like symptoms.

**Key points:**

- Mutations in UBA1 exon 3 have been associated with VEXAS syndrome
- *UBA1* exon 3 was screened in 1650 patients with cytopenia or GCA by amplicon sequencing.
- 6 males were identified from the non-diagnostic cytopenia cohort (1.0%) with *UBA1* mutations.
- A female with a somatic *UBA1* mutation was identified without Monosomy X

## Introduction

Somatic mutations in the gene encoding the major E1 ubiquitin ligase, *UBA1*, cause VEXAS (<u>V</u>acuoles, <u>E</u>1-ligase, <u>X</u>-linked, <u>A</u>uto-inflammatory, <u>S</u>omatic), a late-onset acquired and progressive auto-inflammatory syndrome.[1] Patients with VEXAS are typically male, aged over 60 years with a severe systemic inflammatory disorder, cytopenia and dysplastic features in their bone marrow.[1, 2] Differential diagnoses for patients subsequently found to have VEXAS include, relapsing polychondritis (most frequent diagnosis), Sweet’s syndrome, myelodysplastic syndrome (MDS), giant cell arteritis (GCA) and undifferentiated systemic autoinflammatory disease (uSAID).

To date, cases of VEXAS have been identified by sequencing of exon 3 of *UBA1*, with a particular focus on identifying Methionine-41 (Met41) or splice site mutations.[3] Only 1 amino acid substitution, p.Ser56Phe, has been identified that does not affect Met41, resulting in a temperature dependent reduction of ubiquitin ligase activity.[2] This finding suggests similar missense variants may exist. The milder phenotype observed in the p.Ser56Phe patient indicated individuals with non-Met41 variants may also have milder disease, for which VEXAS may not be considered.

In this study, we sought to investigate the frequency of VEXAS associated mutations in patients with confirmed GCA and those with unexplained cytopenia. This would ultimately inform future diagnostic algorithms for VEXAS screening in patients in either of these scenarios. To achieve this, we developed a high throughput inexpensive assay with which we screened 1,550 samples for all described VEXAS associated *UBA1* mutations identified to date.

## Methods

A one-step, PCR-based amplicon sequencing assay was developed to screen UBA1 in high-throughput (Supplementary Methods). Briefly, each DNA sample was amplified and indexed in a single PCR amplification step by incorporating a unique combination of 8bp DNA barcodes. The resulting amplicons were pooled and sequenced to a minimum depth of 50,000 reads on an Illumina MiSeq. The resulting fastq files were demultiplexed using Flexbar [4], and aligned to the GRCh37 human genome using BWA [5]. Variants were called in gvcf format and genotyping performed per pool (GATK) within a 100bp region encompassing *UBA1* exon 3 and flanking sequence either side (chr.X:47,058,425-47,058,524). Annotation was performed using variant effect predictor (ncbi) and variants identified confirmed by Sanger sequencing using BigDye v3.1 Terminator sequencing kit (Thermo Fisher Scientific), and primers UBA1-Ex3F/3R (Supplementary Table 3).

The cytopenia cohort screened consisted of 1,055 non-diagnostic cases from a large cytopenic cohort (n=2,125), for whom bone marrow DNA was available (Cargo et al, under review). A summary of the cohort can be found in Supplementary Table 1. The cohort includes both male and female adult patients (≥18yrs). All samples were taken with full-informed patient consent for investigation of a suspected haematological disorder, with local ethical committee approval (16/NE/0105).

The GCA cohort screened consisted of 612 males obtained from the UK GCA Consortium (UKGCA)[6, 7]. Favourable ethical opinion was granted by the local Research Ethics Committee (05/Q1108/28) and all participants provided informed written consent. A summary of the cohort can be found in Supplementary Table 2.

## Results and Discussion

1,667 DNA samples were screened for variants in *UBA1* exon 3 by amplicon sequencing based on phenotypes previously associated with VEXAS: cytopenia and GCA. A summary of both cohorts can be found in Table 1. While only male GCA cases were selected, no exclusion criteria were applied to the cytopenia cohort.

**Table 1.** Summary of cohorts screened for *UBA1* mutations by amplicon sequencing. Overview and comparison of the cytopenia and GCA cohorts screened by amplicon sequencing in this study, based on available data. GCA, Giant Cell Arteritis; M/F, Male/Female; CRP, C-Reactive Protein; ESR, Erythrocyte Sedimentation Rate; IQR, Interquartile Range. * Based on n=582/1055 cases with available data, **Based on n=361/612 cases with available data.

| Description | Cytopenia | GCA |
| --- | --- | --- |
| Cohort size | 1055 | 612 |
| Gender (M/F) | 595/460 | 612/0 |
| Median Age [years] (IQR) | 72 (18)* | 71 (10)** |
| Median CRP [mg/L] (IQR) | N/A* | 63 (89)** |
| Median Platelets [$\times 10^9$/L] (IQR) | 115.5 (146)* | 334 (167)** |
| Median ESR [mm/hr] (IQR) | N/A* | 61 (56)** |

In total, 1,650 (98.9%) of samples had a read depth >100 and all 1,667 (100%) had a read depth >30. Despite the more stringent inclusion criteria, no mutations in GCA cases were found, suggesting VEXAS is rarely misdiagnosed as GCA in the UK. In contrast, seven individuals were identified with a *UBA1* mutation (Table 2), in the cytopenic cohort (n=7/1,055, 0.66%). One case (P7) was subsequently found to have been reported in a previous study [1]. All variants identified have been published as a cause of VEXAS and either lead to substitution of Met41 or affect the canonical splice acceptor site. On average, the variant allele frequency (VAF) of the mutation was 44.1% (range:29.5%-59.3%). All variants were confirmed by Sanger sequencing.

**Table 2.** Summary of VEXAS patients identified by amplicon sequencing.

|  | Patient 1 | Patient 2 | Patient 3 | Patient 4 | Patient 5 | Patient 6 | Patient 7 |
| --- | --- | --- | --- | --- | --- | --- | --- |
| **Demographics** |  |  |  |  |  |  |  |
| Deceased | Yes | Yes | Yes | Yes | Yes | Yes | No |
| Gender | M | F | M | M | M | M | M |
| **Investigations** |  |  |  |  |  |  |  |
| Peripheral blood findings Hemoglobin (Hb) g/L, White blood cells (WBC) 10x9/L, Neutrophils (Neu) 10x9/L, Platelets (Plt) 10x9/L | Hb 82, WBC 2.1 neut 0.3, plt 90 | Hb 95, WBC 3.8, plt 98 | Hb 104, WBC 2.63, Neut 0.76, plt 176 | N/A | N/A | Hb 81, WBC 3.8, neut 1.96, plt 211 | HB 103, WBC 5.88, Neu 2.76, Plt 90 |
| Karyotype | Loss of Y in 12/20 cells | N/A | Normal | N/A | N/A | t(8;10) ? Constitutional | Normal |
| Mutations (myeloid panel) | No reportable mutations | No reportable mutations | *ASXL1* c.1771_1772insA p.Tyr591* (VAF 7%) | No reportable mutations | *DNMT3A* c.2160delC p.Lys721Argfs*58 (Vaf 55%) | No reportable mutations | No reportable mutations |
| *UBA1* mutation | c.121A>G, p.Met41Val (VAF 51%) | c.122T>C, p.Met41Thr (VAF 33%) | c.121A>G, p.Met41Val (VAF 41%) | c.121A>G, p.Met41Leu (VAF32%) | c.122T>C, p.Met41Thr (VAF 46%) | c.118-1G>C, p.splice (VAF 19%) | c.122T>C, p.Met41Thr (VAF 52%) |
| **Bone marrow features** |  |  |  |  |  |  |  |
| Cellularity | Increased | Increased | Reduced | No trephine | Increased | Increased | Increased |
| Erythropoiesis | Expanded with minor dysplasia (<10%)occasional vacuoles in erythroblasts. | Minimal dysplasia (<10%) | Minimal dysplasia (<10%) | N/A* | Minimal dysplasia (<10%) occasional vacuolated erythroblasts | minimal dysplasia (<10%) | Reduced |
| Granulopoiesis | Expanded, with minimal dysplasia (<10%). Vacuoles in promyelocytes. | Minimal dysplasia (<10%). Vacuoles in promyelocytes | Minimal dysplasia (<10%). Vacuoles in promyelocytes | N/A* | Expanded, minimal dysplasia (<10%). Vacuoles in promyelocytes | Expanded, minimal dysplasia (<10%). Vacuoles in promyelocytes | Expanded, minimal dysplasia (<10%). |
| Megakaryocytes | Normal number with occasional atypical forms | Normal number with occasional atypical forms | Normal size and morphology | N/A* | Increased with normal morphology | Normal size and morphology | Increased with normal morphology |
| Bone marrow vacuoles | Yes | Yes | Occ | N/A* | Yes | Yes | Yes |
| Bone marrow diagnosis | Reactive | Reactive | Non-diagnostic | Non-diagnostic* | Reactive | No evidence of disease | Non-diagnostic |
| **Inflammatory history** |  |  |  |  |  |  |  |
| Inflammatory features/rheumatological diagnosis | Pulmonary infiltrates, unprovoked DVT, periorbital edema | Fever, WT loss, pulmonary infiltrates, DVT, C-ANCA positive (MPO and PRE negative) vasculitis | unexplained fevers, weight loss, arthritis | Pyoderma gangrenosum-like skin lesion, fevers, uSAID | Not known | Skin rash, fevers non-specific vaculitis | Fevers, Relapsing polychondritis |
| Steroid responsive | Yes | Yes | Not treated | Yes |  | Yes | Yes |

\*Bone marrow sample inadequate for morphological assessment/diagnosis
M-male; F-female; VAF-variant allele frequency; N/A-not available

Six patients had an undiagnosed cytopenia with inflammatory complications, of which five had received steroid treatment and were responsive (Table 2). No clinical information was available for the seventh patient (P5). Re-examination of available bone marrow confirmed the presence of vacuoles in promyelocytes. Importantly these patients had all undergone invasive haematological investigations for unexplained cytopenia without a confirmed diagnosis. Of the seven cases, six of the patients were identified from the 595 male cytopenic cases (1.00%) and had clinical features consistent with previous VEXAS studies (male, over 55 years old with haematological and auto-inflammatory disease).[1, 2, 8] This is the first study investigating the frequency of UBA1 mutations in patients with unexplained cytopenia. The frequency of VEXAS in patients with a confirmed myeloid malignancy has been previously reported by Zhao et al screening 33 males with MDS/CMML for *UBA1* by Sanger sequencing, finding 4 (12%) with mutations.[9] This study was however restricted to male patients and those with confirmed inflammatory/autoimmune disease. Altogether, we add to the growing number of male VEXAS cases with *UBA1* mutations.

Interestingly, one patient (P2) was from the 460 cytopenic female cases (0.02%). Four female VEXAS cases have now been described, all with Monosomy X. [10, 11] We performed a whole genome SNP array on bone marrow DNA from the patient but found no evidence of X chromosome Monosomy or microdeletion around *UBA1*, however we cannot rule out the presence of a smaller (<10kb) deletion. Furthermore, when compared to male VEXAS patients, no difference in variant allele frequency was observed, the VAF of P2 was 33%, within the range observed in the male cases (range: 19%-52%) which may indicate the presence of an undetected deletion. Haematologically, she had minimal dysplasia in her bone marrow cells, increased cellularity and vacuoles in promyelocytes, consistent with male VEXAS cases and displayed similar inflammatory features including fevers and pulmonary infiltrates. To our knowledge, this is the first example of a female with VEXAS without an identified Monosomy X.

Prior to UBA1 screening, all cytopenia patients had undergone prior targeted genetic testing for 27 genes most commonly associated with myeloid malignancy as part of a larger cohort described elsewhere (Cargo et al, under review), as part of their usual clinical care. Two VEXAS cases had a variant in a second gene: P3 was found to harbour a low frequency ASXL1 null mutation (p.Tyr591Ter VAF 7%) and P5 was found to have a *DNMT3A* frameshift mutation (p.Lys721ArgfsTer58, VAF 55%) (Table 2). No studies have yet reported *UBA1* and *ASXL1* variants in the same patient, however somatic *DNMT3A* variants in VEXAS cases have been described.[9, 12] Importantly mutations in these genes, particularly in isolation, have been reported in aging healthy individuals [13] and therefore could represent coincidental age-related clonal haematopoiesis. Ultimately the significance of these co-occurring variants in the same patient remains unknown.

In summary we describe a high-throughput screen for *UBA1* exon 3 and identify seven cases of VEXAS syndrome in patients with unexplained cytopenia, including one female patient without Monosomy X. Screening for *UBA1* mutations in patients with unexplained cytopenia should therefore be considered, particularly in those presenting with inflammatory or autoimmune conditions. Our amplicon assay provides a high-throughput inexpensive method to screen large cohorts and inclusion of *UBA1* in myeloid targeted sequencing panels should also be considered.

## Data Availability

All data produced in the present work are contained in the manuscript. Sequence data have been submitted and are available from NCBI Sequence Read Archive (ref. PRJNA784094).

## Acknowledgements

J.A.P is supported by a UKRI Future Leaders Fellowship (MR/T02044X/1). GCA research is also supported by an MRC Treatment According to Response in Giant cEll arteritis (TARGET) Partnership award to AWM and is supported by the National Institute for Health Research (NIHR) Leeds Biomedical Research Centre and a NIHR Senior Investigator award to AWM. The views expressed are those of the author(s) and not necessarily those of the NHS, the NIHR or the Department of Health. This study was partially funded by unrestricted research grant from SOBI pharmaceuticals.

The members of the UKGCA Consortium include Andrew Gough, John D. Isaacs, Michael Green, Neil McHugh, Lesley Hordon, Sanjeet Kamath, Mohammed Nisar, Yusuf Patel, Cee-Seng Yee, Robert Stevens, Pradip Nandi, Anupama Nandagudi, Stephen Jarrett, Charles Li, Sarah Levy, Susan Mollan, Abdel Salih, Oliver Wordsworth, Emma Sanders, Esme Roads, Anne Gill, Lisa Carr, Christine Routledge, Karen Culfear, AsankaNugaliyadde, Lynne James, Jenny Spimpolo, Andy Kempa, Felicity Mackenzie, Rosanna Fong, Genessa Peters, Bridie Rowbotham, Zahira Masqood, Jane Hollywood, Prisca Gondo, Rose Wood, Steve Martin, Lubna Haroon Rashid, James I. Robinson, Mike Morgan, Louise Sorensen, and John Taylor.

## Author Contributions

JP, SS and CC designed the study. JP performed the sequencing and analysis of DNA samples. CC, PE, SM, SLM and AWM recruited patients and collected DNA samples. MG, BA, EN, DIDT and CC provided associated clinical data. CC and AK performed bone marrow/aspirate analysis. JP and SS wrote the first draft of the manuscript. All authors read and approved the manuscript.

## Conflict of Interest Disclosures

CC has contributed to advisory boards for Novartis and AOP Orphan and has research funding from Celgene. PE has received honoraria from Astellas. The remaining authors declare no conflicts of interest.

## Notes

### Author Declarations

All cytopenia samples were taken with full-informed patient consent for investigation of a suspected haematological disorder. The North East - York Research Ethics Committee gave ethical approval for this work (REC ref: 16/NE/0105). For the GCA cohort, the Leeds West Research Ethics Committee gave ethical approval for this work (REC ref: 05/Q1108/28).

