## Supplementary Data for "A high-throughput amplicon screen for somatic *UBA1* variants in Cytopenic and Giant Cell Arteritis cohorts"

### Supplementary Tables

Supplementary Table 1 – Summary of cytopenia cohort
Supplementary Table 2 – Summary of GCA cohort
Supplementary Table 3 – Primers used for amplicon sequencing

### Supplementary Methods

*UBA1* exon 3 pooled amplicon sequencing

**Supplementary Table 1 - Summary of cytopenia cohort**

<u>Age (n=1055)</u>

| | |
|---|---|
| **Mean** | 68.0 |
| **Median** | 72 |
| **Range** | 18-94 |

<u>Gender</u>

| | |
|---|---|
| **Males** | 595 |
| **Females** | 459 |

**Whole Cohort (n=1055)**

| | wbc | rbc | hb | hct | mcv | plt | lymph % | mixed % | neutr % |
|---|---|---|---|---|---|---|---|---|---|
| **Mean** | 5.53 | 4.35 | 113.63 | 33.64 | 91.02 | 148.43 | 28.86 | 13.84 | 58.08 |
| **Median** | 4.8 | 3.69 | 113 | 33 | 90.2 | 115.5 | 26.1 | 12.4 | 60.7 |
| **Range** | 0.9-21.4 | 1.52-141 | 33.9-174 | 16.9-97.7 | 59.8-182 | 0-764 | 1-78.3 | 1.1-51.2 | 12.5-95.5 |
| **Missing data** | 473 | 473 | 473 | 473 | 473 | 471 | 473 | 473 | 473 |

*wbc, white blood count; rbc, red blood count; hb, haemoglobin; hct, ; mcv, ; plt, platelets; lymph%, percentage lymphoid, mixed %, ; neutr %, percentage of neutrophils*

**Supplementary Table 2 - Summary of GCA cohort**

Age (n=361 with available data)

| | |
|---|---|
| **Median** | 71 |
| **IQR** | 10 |

Gender (n=361 with available data)

| | |
|---|---|
| **Males** | 612 |
| **Females** | 0 |

Blood Biochemistry (at presentation)

| | n = | Median | IQR |
|---|---|---|---|
| ESR (mm/hr) | 361 | 61 | 56 |
| CRP (mg/L) | 361 | 63 | 89 |
| Platelets (x10^9/L) | 361 | 334 | 167 |
| Missing Data | 251 | | |

Reported phenotypes associated with GCA

| | yes | no | unknown |
|---|---|---|---|
| Neoplasia | 33 | 338 | 241 |
| PMR symptoms | 181 | 376 | 55 |
| Ischemic complications | 30 | 313 | 269 |

**Supplementary Table 3 – Primers used for amplicon sequencing**

| Primer Name | Sequence 5'-3' (sequencing adapter – index – *UBA1* primer) |
|---|---|
| UBA1-i7_A-F | ACACTCTTTCCCTACACGACGCTCTTCCGATCT-GTGAATAT-TGTCTGTCTATCCATGCTCCAC |
| UBA1-i7_B-F | ACACTCTTTCCCTACACGACGCTCTTCCGATCT-ACAGGCGC-TGTCTGTCTATCCATGCTCCAC |
| UBA1-i7_C-F | ACACTCTTTCCCTACACGACGCTCTTCCGATCT-CATAGAGT-TGTCTGTCTATCCATGCTCCAC |
| UBA1-i7_D-F | ACACTCTTTCCCTACACGACGCTCTTCCGATCT-TGCGAGAC-TGTCTGTCTATCCATGCTCCAC |
| UBA1-i7_E-F | ACACTCTTTCCCTACACGACGCTCTTCCGATCT-TCTCTACT-TGTCTGTCTATCCATGCTCCAC |
| UBA1-i7_F-F | ACACTCTTTCCCTACACGACGCTCTTCCGATCT-CTCTCGTC-TGTCTGTCTATCCATGCTCCAC |
| UBA1-i7_G-F | ACACTCTTTCCCTACACGACGCTCTTCCGATCT-CCAAGTCT-TGTCTGTCTATCCATGCTCCAC |
| UBA1-i7_H-F | ACACTCTTTCCCTACACGACGCTCTTCCGATCT-TTGGACTC-TGTCTGTCTATCCATGCTCCAC |
| UBA1-i5_1-R | GACTGGAGTTCAGACGTGTGCTCTTCCGATCT-CTAGCGCT-CCACCCAACCTTATCCTCTC |
| UBA1-i5_2-R | GACTGGAGTTCAGACGTGTGCTCTTCCGATCT-TCGATATC-CCACCCAACCTTATCCTCTC |
| UBA1-i5_3-R | GACTGGAGTTCAGACGTGTGCTCTTCCGATCT-CGTCTGCG-CCACCCAACCTTATCCTCTC |
| UBA1-i5_4-R | GACTGGAGTTCAGACGTGTGCTCTTCCGATCT-TACTCATA-CCACCCAACCTTATCCTCTC |
| UBA1-i5_5-R | GACTGGAGTTCAGACGTGTGCTCTTCCGATCT-CGCTATGT-CCACCCAACCTTATCCTCTC |
| UBA1-i5_6-R | GACTGGAGTTCAGACGTGTGCTCTTCCGATCT-TATCGCAC-CCACCCAACCTTATCCTCTC |
| UBA1-i5_7-R | GACTGGAGTTCAGACGTGTGCTCTTCCGATCT-TCTGTTGG-CCACCCAACCTTATCCTCTC |
| UBA1-i5_8-R | GACTGGAGTTCAGACGTGTGCTCTTCCGATCT-CTCACCAA-CCACCCAACCTTATCCTCTC |
| UBA1-i5_9-R | GACTGGAGTTCAGACGTGTGCTCTTCCGATCT-CCGCGGTT-CCACCCAACCTTATCCTCTC |
| UBA1-i5_10-R | GACTGGAGTTCAGACGTGTGCTCTTCCGATCT-TTATAACC-CCACCCAACCTTATCCTCTC |
| UBA1-i5_11-R | GACTGGAGTTCAGACGTGTGCTCTTCCGATCT-CAAGCTAG-CCACCCAACCTTATCCTCTC |
| UBA1-i5_12-R | GACTGGAGTTCAGACGTGTGCTCTTCCGATCT-TGGATCGA-CCACCCAACCTTATCCTCTC |
| UBA1-3F | TGTCTGTCTATCCATGCTCCAC |
| UBA1-3R | CCACCCAACCTTATCCTCTC |

**Supplementary Methods**

UBA1 exon 3 pooled amplicon sequencing

Primers containing a *UBA1* priming sequence, unique 8bp index and partial Illumina sequencing adapters (Supplementary Table 3) were manufactured by Integrated DNA Technologies (Leuven, Belgium). Unique combinations of 8 forward and 12 reverse primers were pooled in a 96 well plate to allow demultiplexing of sequenced data. 50-100ng of genomic DNA was amplified in 96 well plates using High-Fidelity Phusion 2x Mastermix (Thermo Fisher Scientific) and 5uM of each primer pair. Following amplification, 2ul of each PCR product was pooled and an AMPure bead (Beckman Coulter, High Wycombe, UK) clean-up performed using a 1:1 volume ratio of AMPure beads:DNA. 25ul of each pool at a concentration of 20ng/ul was sequenced using a 250bp paired-end read protocol on an Illumina MiSeq (Genewiz, Boston, USA).
